# The Pan-Cancer Landscape of Prognostic Germline Variants in 10,582 Patients

**DOI:** 10.1101/19010264

**Authors:** Ajay Chatrath, Roza Przanowska, Shashi Kiran, Zhangli Su, Shekhar Saha, Briana Wilson, Takaaki Tsunematsu, Ji-Hye Ahn, Kyung Yong Lee, Teressa Paulsen, Ewelina Sobierajska, Manjari Kiran, Xiwei Tang, Tianxi Li, Pankaj Kumar, Aakrosh Ratan, Anindya Dutta

## Abstract

While clinical data provides physicians with information about patient prognosis, genomic data can further improve these predictions. We analyzed sequencing data from over 10,000 cancer patients and identified hundreds of prognostic germline variants using multivariate Cox regression models. These variants provide information about patient outcomes beyond clinical information currently in use and may augment clinical decisions based on expected tumor aggressiveness. Molecularly, at least twelve of the germline variants are likely associated with patient outcome through perturbation of protein structure and at least five through association with gene expression differences. About half of these germline variants are in previously reported tumor suppressors or oncogenes, with the other half pointing to loci of previously unstudied genes in the literature that should be further investigated for roles in cancers. Our results suggest that germline variation contributes to tumor progression across most cancers and contains patient outcome information not captured by clinical factors.

## Introduction

Large-scale sequencing projects^1^ increased our molecular understanding of cancers^2^ to the point where using sequencing data to augment clinical decisions seems promising. Somatic mutations in cancers have received substantial attention in oncology^2^ as they can be used to individualize drug selection.^3^ While much effort has been directed towards characterizing somatic mutations in cancer, recent studies suggest that germline variants also have significant clinical utility.

In line with the heritability of some cancers,^4^ several germline variants predict a patient’s risk for developing cancer and are useful for individualizing cancer screening guidelines.^5-13^ Germline variation can affect drug sensitivity,^14-20^ predict drug toxicity, and could help select therapy to minimize side-effects.^21-26^ Some germline variants increase patient risk for specific somatic aberrations, suggesting that germline variation may impact disease course.^27^

We hypothesized that the effects of germline variants on cancer progression may be strong enough to identify associations with patient outcome. Previous studies tested for an association between patient outcome and a small number of germline variants in genes well-characterized in a given cancer.^28,29^ We published an unbiased method of testing for an association between a large number of germline variants and patient outcome in patients with lower grade gliomas.^30^ In this study, we identify prognostic germline variants using sequencing data from 10,582 patients from The Cancer Genome Atlas (TCGA). These germline variants significantly improve predictions of patient outcome compared to clinical variables alone, identify biological mechanisms by which germline variants affect patient outcomes, and identify genes and pathways that impact cancer biology and therapy.

## Results

### Identification of High Quality Germline Variants

Germline variants were called and filtered as shown in **eFigure 1** using sequencing data from 10,582 TCGA patients with 33 different types of cancers. In total, 77.6 million unique variants were called. After filtering, we limited our analysis to 519,319 unique variants (**eFigure 2**). Because the final variant call set was created by merging variant calls from whole exome sequenced (WXS) normal tissue samples, WXS tumor samples, and RNA sequenced tumor samples, we evaluated our variant calls for contamination by somatic mutations or RNA editing. Our final germline variant call set did not substantially overlap with somatic mutations or RNA editing sites (**eFigures 3-4, eText 1**).

### Determination of Prognostic Clinical Models for Each Cancer

To identify prognostic germline variants that provide additional outcome information not already captured by clinical variables, we created clinical models predictive of patient outcome for each cancer using the clinical information previously collected by the TCGA research network along with calculated race. The variables selected for each cancer are summarized in **eTable 1**. The study was powered to capture prognostic germline variants with moderate to high effect sizes (beginning at hazard ratios > 2) (**eFigure 5, eText 2**).

### Identification of Prognostic Germline Variants

The 191 prognostic variants from the six analyses are described in **eTable 2A-F**.

The first three analyses identified germline variants associated with prognosis in (1) individual cancers, (2) multiple cancers giving roughly equal weight to each cancer, and (3) cancers grouped by organ system, histological, or molecular classifications (**Figure 1A**). Analysis 1 tested 519,139 variants for associations with patient outcome in individual cancers and identified 70 unique prognostic variants (**Figure 1B, eTable 2A**, Kaplan Meier plots of selected examples in **Figure 2**).

**Figure 1.**
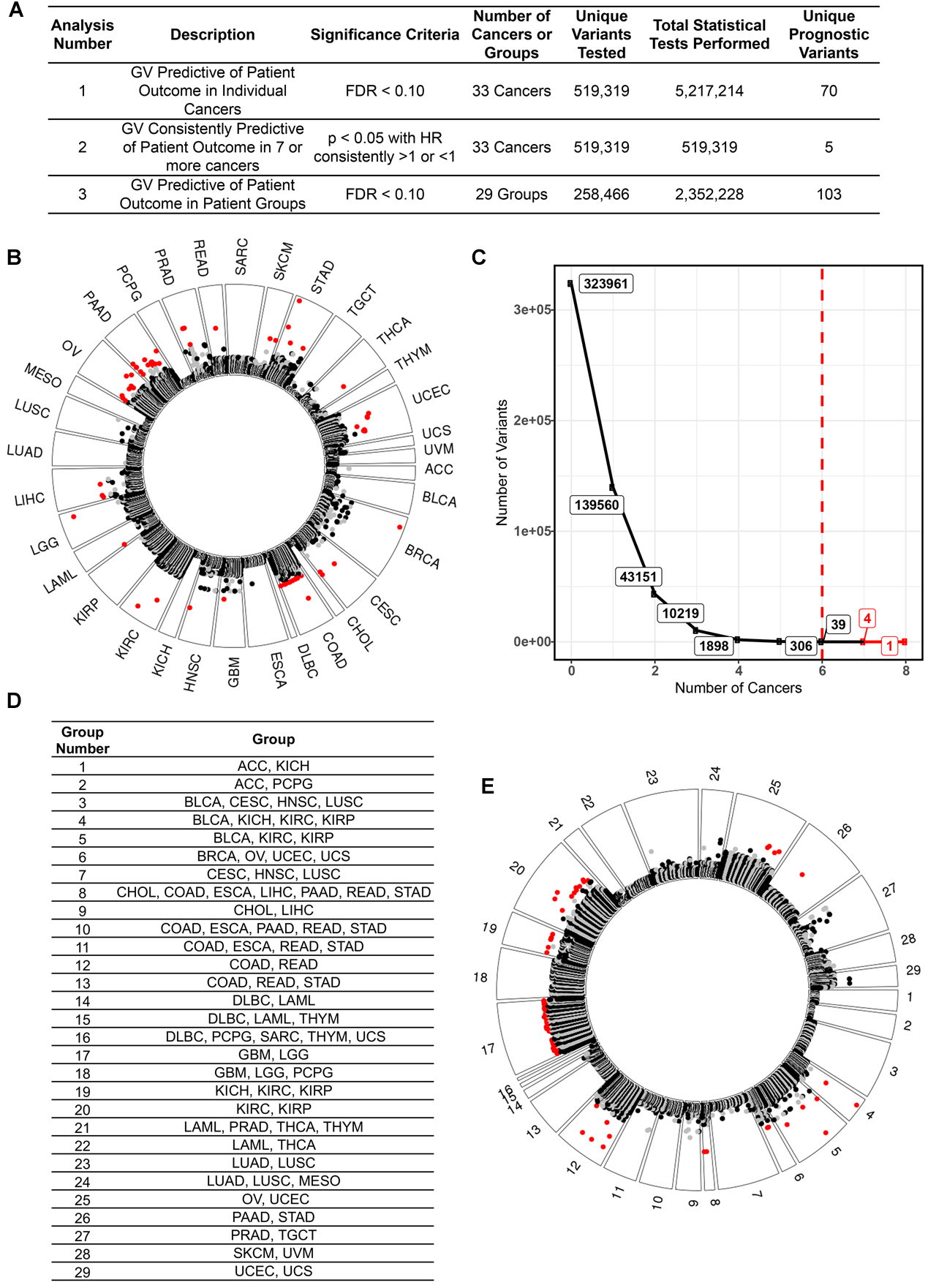
Prognostic germline variants identified in analyses one through three. **A**. A description of the three analyses used to identify prognostic germline variants in this figure. **B**. Analysis 1. Germline variants found to be predictive of patient outcome in each cancer. Each dot represents a germline variant that was tested for an association with patient outcome. Variants closer to the outside of the plot are more closely associated with patient outcome. Variants in red are significantly (FDR<0.10) associated with patient outcome. The alternating black and grey colors reflect alternating chromosomes for the germline variants that were not significant predictors of patient outcome. **C**. Analysis 2. Germline variants found to be recurrently predictive of patient outcome in multiple different cancers. We identified 5 total germline variants that were recurrently predictive (p<0.05) of favorable (HR<1) or deleterious (HR>1) patient outcomes in 7 or more different cancers. **D**. Analysis 3. 29 groups of cancers created to identify germline variants with weaker effect sizes in larger patient cohorts. Justification for these groups is provided in **eTable 3**. **E**. Analysis 3. Germline variants found to be predictive of patient outcome in the groups described in **Figure 1D**. The format of the figure is the same as in **Figure 1B**.

**Figure 2.**
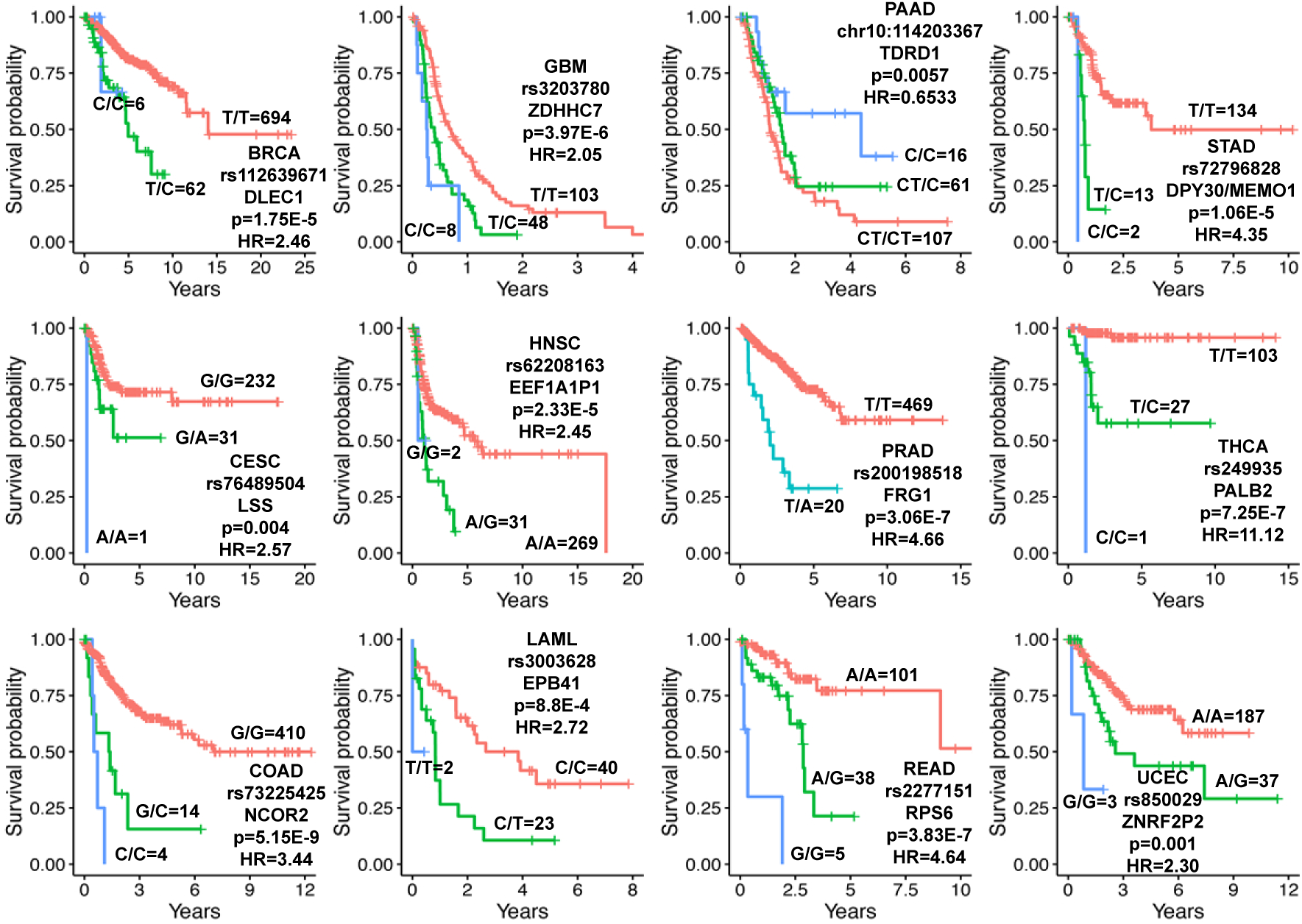
Selected Kaplan-Meier plots of the prognostic germline variants from Analysis 1. The number of patients in each group is indicated next to each line and the patient outcome measure of each disease is given in **eTable 1**. The reported p-values and hazard ratios were calculated using univariate regression and are different from the p-values and hazard ratios reported elsewhere which are based on multivariate regression.

While analysis 2 identified hundreds of variants recurrently predictive of outcome in >4 cancers, we will only discuss the 5 variants that were predictive in seven or more cancers (**Figure 1C, eTable 2B**). Both the direction of the hazard ratios (deleterious or protective) and the magnitude of the effect on patient outcome for germline variants across different cancers were highly correlated (**eText 3**).

Analysis 3 increased our statistical power by grouping similar cancer types to increase the number of patients with the minor allele that could be included in the study. 29 different patient groups were created based on organ system, histological, or molecular classification (**Figure 1D**, group justification in **eTable 3**). 258,466 unique germline variants were tested and 103 prognostic variants were identified (**Figure 1E, eTable 2C**, Kaplan Meier plots of selected examples in **eFigure 6**).

### Prognostic Germline Variants Causing Significant Amino Acid Changes

Analyses 4-6 repeated analyses 1-3 but limited these analyses to variants within the top 0.3% of deleterious mutants across the human genome with CADD>25 (**Figure 3A**). Analysis 4 tested a total of 981 unique variants and identified nine unique prognostic variants (**Figure 3B, eTable 2D**). Of the 16 variants that were recurrently predictive of patient outcomes in 4 or more cancers (analysis 5), we will discuss the one variant that was predictive in five cancers (**Figure 3C, eTable 2E**). Analysis 6 tested 903 unique variants for an association with outcome in the patient groups used in analysis 3 and described in **Figure 1D** and identified 3 additional prognostic variants (**Figure 3D, eTable 2F**).

**Figure 3.**
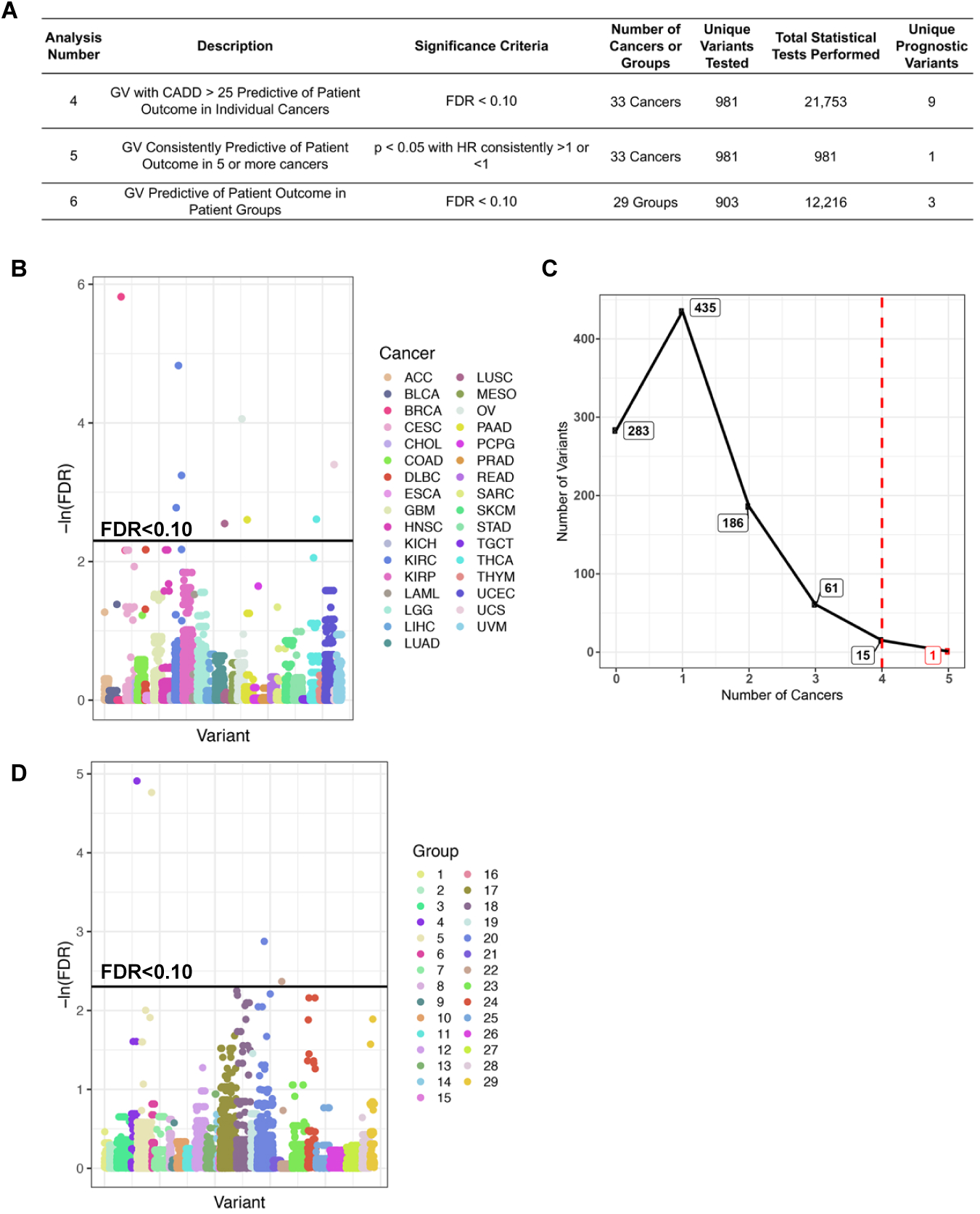
Prognostic germline variants that cause significant amino acid changes (CADD>25) identified in analyses four through six. **A**. A description of the three analyses used to identify prognostic germline variants in this figure. **B**. Analysis 4. Germline variants causing significant amino acid changes found to be predictive (FDR<0.10) of patient outcome in each cancer. **C**. Analysis 5. Germline variants causing significant amino acid changes found to be recurrently predictive (p<0.05) of favorable (HR<1) or poor (HR>1) patient outcomes in 5 or more different cancers. **D**. Analysis 6. Germline variants causing significant amino acid changes found to be predictive of patient outcome in patient groups defined in **Figure 1D**.

### The Pan-Cancer Landscape of Prognostic Germline Variants

The large number of prognostic variants identified in analysis 1 and 3 allowed us to compare the characteristics of these germline variants with previously reported characteristics of variants identified by genome wide association studies (GWAS). Three characteristics have been noted in variants identified through GWAS: (1) the minor allele tends to be deleterious when considering the set of variants with large effect sizes, (2) there is a negative correlation between effect size and allele frequency, and (3) most germline variants identified by GWAS do not cause amino acid changes.^31^

To test whether the deleterious allele is usually the minor allele, the predictive alternate alleles from analysis 1 were classified as deleterious (HR>1) or protective (HR<1) based on the Cox regression results. Of the prognostic germline variants from analysis 1, the deleterious allele is clearly often the minor allele (p=7.077E-8) (**Figure 4A**). A similar analysis with the predictive variants from analysis 3 (**Figure 4B**) did not show a significant statistical depletion of deleterious alternate alleles from the population (p=0.115). The predictive variants from analysis 3 were detectable only with larger sample sizes and have smaller effect sizes than those identified by analysis 1. Thus the result in **Figure 4B** is still consistent with the first premise^31^ that a deleterious allele with a large effect size (as in analysis 1, but not analysis 3) is usually the minor allele.

**Figure 4.**
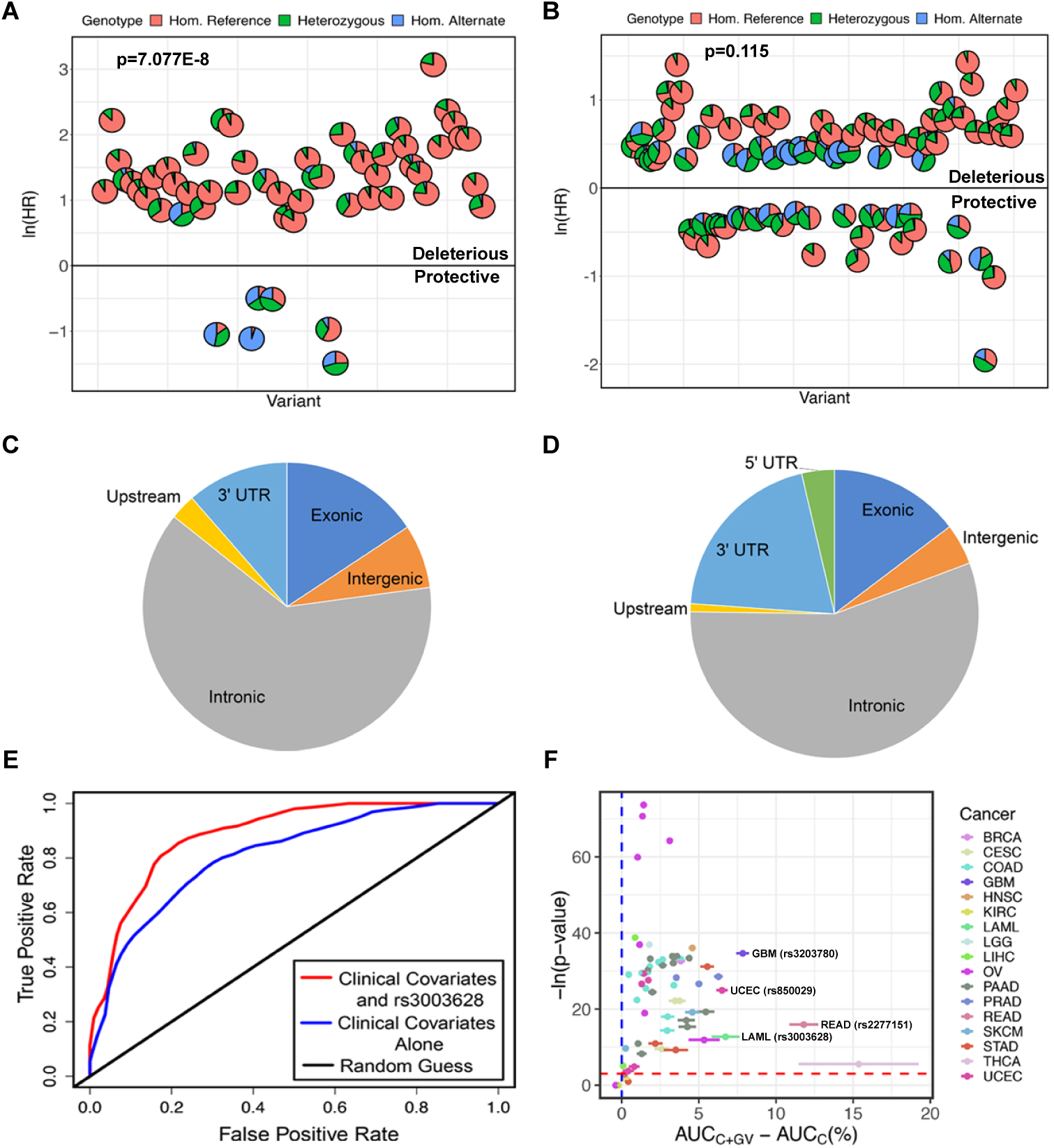
Characteristics of prognostic germline variants and improvement of patient outcome models by the prognostic germline variants. **A-B**. Scatterplots of the prognostic germline variants identified in individual cancers in Analysis 1 (**A**) and in groups of cancers in Analysis 3 (**B**). Each pie chart reflects the distribution of patients that are homozygous for the reference allele, heterozygous, and homozygous for the alternate allele for one prognostic variant. The minor allele was much more likely to be deleterious than protective (p=7.077E-8) in Analysis 1 though this trend was not significant in Analysis 3 (p=0.115). **C-D**. Pie charts displaying the genomic locations of the germline variants in Analysis 1 (**C**) and Analysis 3 (**D**). **E**. An example of a receiver operator characteristic (ROC) curve calculated using data from LAML at 366 days of follow-up. The blue line represents the patient outcome predictions made using clinical information alone (C model). The red line represents patient outcome predictions made using clinical information in addition to rs3003628 germline variant status (C+GV model), which we found to be predictive of patient outcomes in LAML. The Area Under the Curve (AUC) was 0.81 for the C model and 0.93 for the C+GV model giving a ΔAUC of 0.12 (12%). **F**. Many of the prognostic germline variants improve clinical outcome model predictions. For each prognostic variant, we created a ROC curve based on the clinical (C) model and the clinical + germline variant (C+GV model), as in **Figure 5E**, at each point in time from the 10^th^-90^th^ percentile of patient progression or death for each cancer. The ΔAUC of the C+GV model versus the C model at each time point was calculated (**eTable 4**). X-axis: Mean and standard error of ΔAUC. Y-axis: The p-values from testing whether or not the AUC of the C+GV model is significantly greater than that of the C model using a Wilcoxon rank sum test. Four examples of prognostic germline variants that significantly increase the AUC are labeled and highlighted in **eTable 4**.

A negative correlation is seen between effect size and allele frequency with both variants from analysis 1 (Spearman’s rho = -0.282, p=0.0184) and analysis 3 (Spearman’s rho = -0.667, p<2.2E-16), satisfying the second premise. Finally, the vast majority of predictive variants identified by this GWAS do not cause amino acid changes (**Figure 4C-D**), satisfying the third premise.

A previous study had identified germline variants associated with an increased incidence of somatic mutations in cancer related genes.^27^ We also find that somatic mutations are increased in driver genes in the relevant cancer with prognostic versus non-prognostic variants (OR=1.89, p=0.0001, **eText 4**).

### Germline Variants Significantly Improve Outcome Prediction Models

The effect sizes of prognostic germline variants from analysis 1 were large enough to hypothesize that germline variants identified in individual cancers could improve clinical outcome models in current use.

The clinical variables predictive of outcome (**eTable 1**) were used to generate the first outcome model (Clinical: C). The second outcome model was based on clinical information plus the status of a particular predictive germline variant (Germline Variant: GV) (C+GV). An example receiver operator characteristic (ROC) curve for predicting LAML patient vital status at 366 days of follow-up is shown using C and C+GV for predictive variant rs3003628 (ROC in **Figure 4E**). The area under the ROC curves (AUC) for the C model is 0.807 and for the C+GV model is 0.928. The change in AUC (ΔAUC) for the C+GV model relative to the C model in this example is 0.12 (12%). To ensure that the change in AUC is consistent at different times of follow-up, ΔAUC was calculated from the 10^th^ to the 90^th^ percentile of patient outcome time. The mean and standard error of ΔAUC was plotted against the p-value of the one-sided test evaluating whether the AUC for C+GV is significantly larger than the AUC for C (**Figure 4F**).

This analysis was repeated for all predictive variants. There is a consistent, statistically significant (p<0.05) increase in AUC when the clinical model is enhanced by germline variant information (C+GV) compared to the clinical model alone (C) for 63 of the predictive germline variants out of 70 tested (**eTable 4**). These results demonstrate that adding predictive germline variants to existing clinical criteria will improve the prediction of outcome of many cancers.

### Prognostic Variants in Driver Genes, Oncogenes, and Tumor Suppressor Genes

90 of the 193 genes in the proximity of one of the prognostic germline variants have been functionally implicated in nine of the twelve hallmarks of cancer (**Figure 5A, eTable 5**).^32^

**Figure 5.**
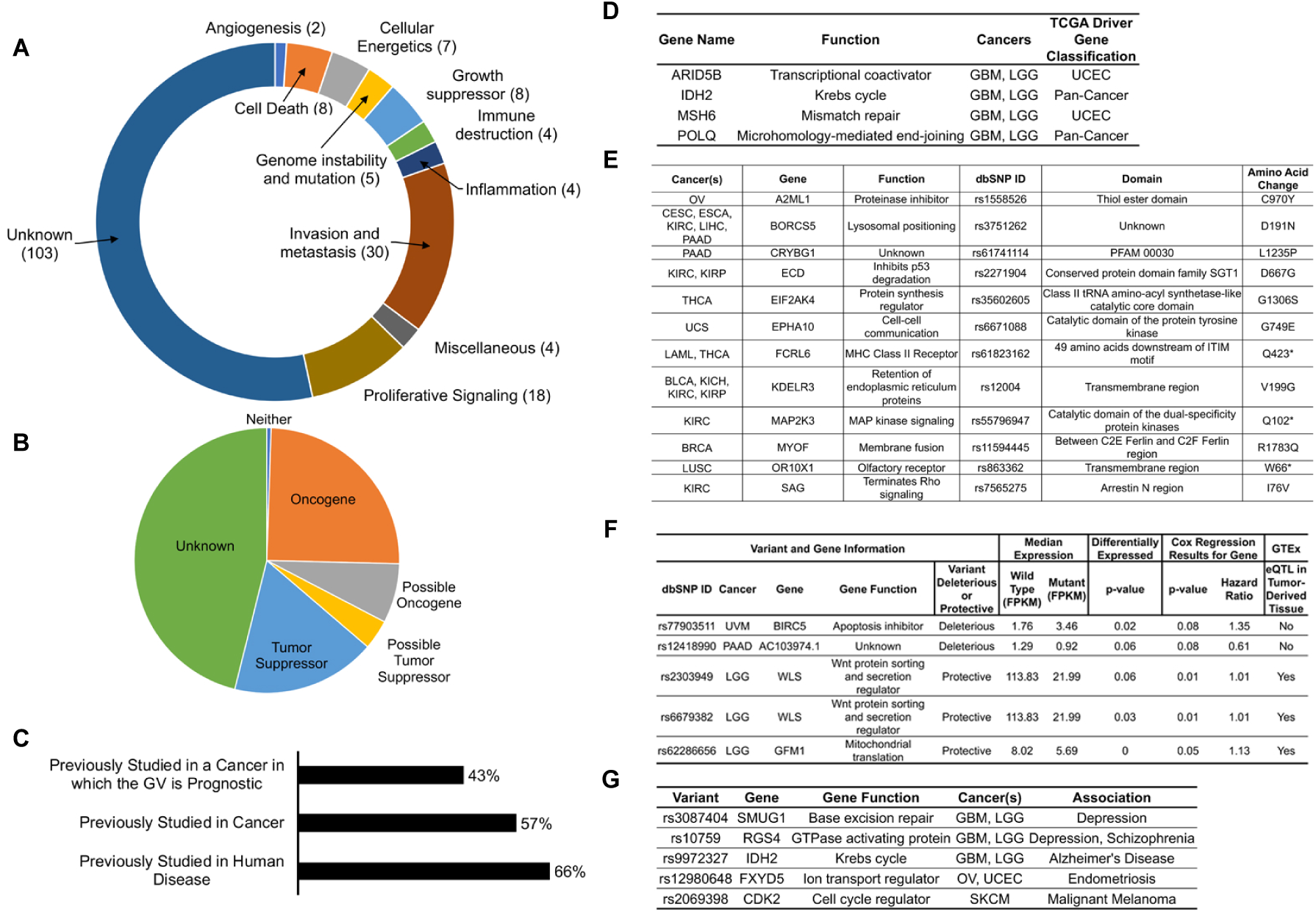
Literature review of genes associated with the prognostic germline variants and mechanisms by which prognostic germline variants may exert their effects. **A**. The cancer-related functions of genes associated with the prognostic germline variants are quite diverse. **B**. Many of the genes associated with the variants have previously been reported to be tumor suppressor genes or oncogenes. We categorized genes as tumor suppressor genes or oncogenes based on phenotypes reported in the literature, even if the exact mechanism through which the genes act have not yet been determined. **C**. Although many of the variants have been studied in the field, there are many genes that have not yet been studied in the context of human disease and therefore may warrant investigation by the field. **D**. Four of the genes associated with prognostic germline variants are in previously reported cancer driver genes. **E**. Some of the prognostic germline variants cause dramatic amino acid changes and may disrupt well-characterized protein domains. **F**. Some of the prognostic germline variants likely act as expression quantitative trait loci in *cis* (*cis* eQTLs) and the expression of these genes are predictive of patient outcome. We found three of these germline variants to also be eQTLs in the genotype tissue expression (GTEx) database in the same tissue that the tumor was derived from. **G**. Some of the prognostic germline variants have been reported to be associated with other diseases related to the tissue from which the tumor was derived.

Roughly 50% of the predictive variants are found in or near genes that possibly have tumor suppressor or oncogenic activity (**Figure 5B, eTable 5**). About 25% of the predictive genes were previously studied in the cancer in which the germline variant was found to be prognostic, about half were previously studied in at least one cancer, and roughly two-thirds were studied in at least one human disease (**Figure 5C, eTable 5**). Prognostic variants were identified in or near *MSH6, POLQ, ARID5B*, and *IDH2*, which are previously reported cancer-driver genes (**Figure 5D**).

### Prognostic Germline Variants Can Cause Significant Amino Acid Changes or Act as eQTLs

The 12 prognostic variants identified in analyses 4-6 caused significant amino acid changes (CADD>25), with many of these amino acid changes occurring in protein-coding domains with annotated or known functions (**Figure 5E**).

39 variants could act as *cis* eQTLs, as they were associated with expression differences of the proximate genes. We highlight 5 of these variants because the expression levels of the proximate genes are also predictive of survival, with the direction of the effect (HR >1 or <1) being concordant with the effect of the variant (**Figure 5F**). Of these 5 variants, 3 were also *cis* eQTLs in the corresponding tissue in GTEx.^33^

### Prognostic Variants Implicated in Other Diseases

Some of the prognostic variants are linked with diseases that occur in the tissue giving rise to the tumor, suggesting the variant has an important function in that tissue (**Figure 5G, eTable 6A**). **eTable 6B** lists prognostic genes that are linked in the literature to traits in tissues outside the ones bearing the tumors.

### Individual Prognostic Variant Characterization

In this section, we characterize three germline variants to illustrate how individual germline variants may be associated with patient outcome.

### rs1800932 in *MSH6* May Be Associated with Favorable Outcome by Increasing Temozolomide Sensitivity

rs1800932 predicts favorable patient outcome in gliomas (LGG and GBM). This variant is an eQTL for increased expression of *MSH6* in many tissues, including nerve,^33^ is associated with increased expression of *MSH6* in patients with LGG (p=0.00732), and has previously been reported to be associated with a decreased risk of prostate cancer.^34^ We found *MSH6* expression to be correlated with elevated temozolomide sensitivity in cancer cell lines (Spearman’s rho=0.165, p=5.01E-7).^35^ Temozolomide is a DNA alkylating agent used in the treatment of most glioma patients and is likely to have been used in the therapy of the most patients with gliomas in TCGA. *MSH6* knockdown increases temozolomide resistance^36^ and somatic mutations in *MSH6* are associated with temozolomide resistance in gliomas.^37^ Taken together, this suggests that rs1800932 is an eQTL for increased expression of *MSH6* in gliomas, which may increase sensitivity to temozolomide, the primary chemotherapeutic agent for gliomas.

### rs55796947 in *MAP2K3* May Result in Cell Cycle Arrest and Apoptosis

rs55796947 in *MAP2K3/MKK3* predicts favorable prognosis in KIRC. This germline variant introduces a stop codon in *MAP2K3* that truncates the kinase domain. *MAP2K3* inhibition results in cell cycle arrest, autophagy-mediated cell death, the unfolded protein response (UPR), and sensitization to chemotherapy drugs.^38^ Indeed, tumors in patients with this variant upregulate genes involved with apoptosis (p<0.001, **Figure 6A-B**) and downregulate *E2F* targets involved in cell-cycle progression (p=0.047, **Figure 6C**). This germline variant likely truncates the kinase domain of *MAP2K3*, resulting in cell cycle arrest, apoptosis, and favorable patient outcome.

**Figure 6.**
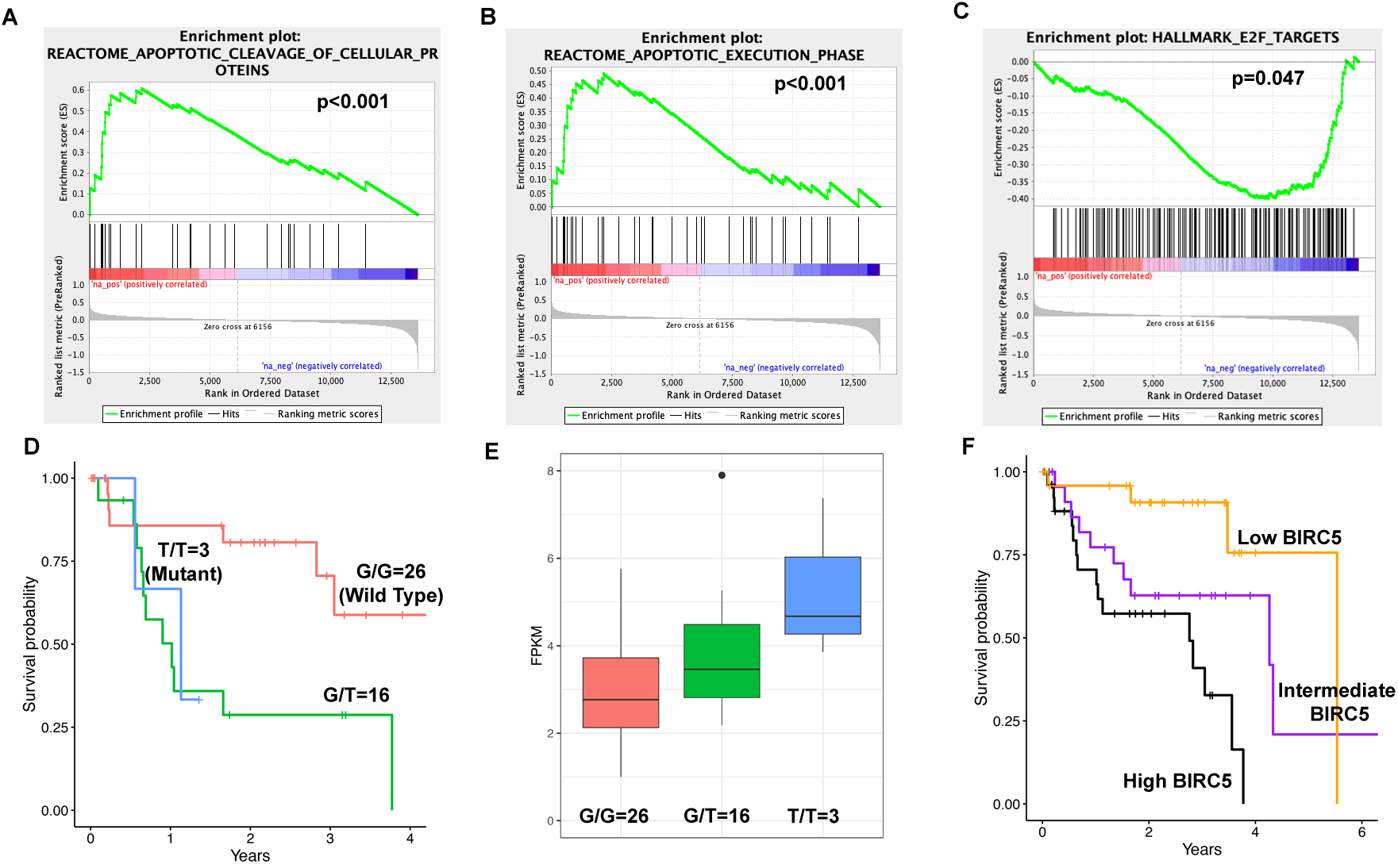
Examples by which two of the prognostic germline variants may be associated with patient outcome. **A-C**. rs55796947 in *MAP2K3/MKK3* is associated with favorable patient outcome in KIRC and results in complete loss of *MAP2K3*’s protein kinase domain due to a Q73* amino acid change. *MAP2K3* inhibition has previously been reported to result in cell cycle arrest and response to chemotherapy drugs. Tumors with the variant show upregulation of genes involved with apoptotic cleavage (**A**), genes in the apoptotic execution phase (**B**), and downregulation of E2F targets (**C**) in a Gene Set Enrichment Analysis (GSEA) of RNAseq data. **D-F**. rs77903511 in the apoptosis inhibitor *BIRC5* is predictive of poor patient outcome in UVM (**D**). This variant is associated with increased *BIRC5* expression (**E**). Elevated *BIRC5* expression is associated with poor patient outcome (**F**).

### rs77903511 is an eQTL for *BIRC5* which Inhibits Apoptosis

rs77903511 predicts poor patient outcome in UVM (**Figure 6D)**. *BIRC5* inhibits apoptosis through interaction with and inhibition of caspase 9 and effector caspases. The alternate allele is associated with increased *BIRC5* expression in the tumors (p=0.02, **Figure 6E**). Consistent with a role of *BIRC5* in apoptosis inhibition, *BIRC5* expression is associated with poor patient outcome (**Figure 6F**). This variant, therefore, may be associated with poor outcome because of an increase of the apoptosis inhibitor *BIRC5*.

## Discussion

This study shows, as a general principle, that germline variants are associated with cancer patient outcome. The prognostic germline variants enhanced patient outcome predictions compared to models based on currently collected clinical data. We envision germline variants providing clinicians with information about a patient as a supplement to reported history, physical exam findings, and imaging and laboratory tests. These predictions will improve over time with the use of more information available in electronic medical records. While we identified a large number of prognostic germline variants in analysis 1, the power calculations and the identification of additional prognostic germline variants by grouping similar cancers suggests that more prognostic germline variants will emerge as more tumors are sequenced.

Further study is necessary to validate the associations that we identified, as setting the discovery threshold at FDR<0.10 suggests that some of the associations may have occurred by random chance. The variants identified in analyses 2 and 5 require deeper interrogation, as we were unable to develop an unbiased test to assess the probability of those associations occurring by random chance. While we identified germline variants associated with significant improvements in clinical outcome predictions, further work is necessary to identify situations in which the additional prognostic information would be valuable for treatment decisions or end of life planning.

It is reassuring that a significant fraction of prognostic germline variants are found in or near possible tumor suppressor genes, oncogenes, or known cancer driver genes. The variants in cancer driver genes, *MSH6, POLQ, ARID5B*, and *IDH2*, warrant further study to determine the mechanism by which these variant affect cancer progression.^39^ The twelve germline variants in **Figure 5E** that cause substantial amino acid changes are prime candidates for experimental follow-up and are discussed in detail in **eText 5**. A handful of the prognostic germline variants have been associated with human disease, some in the same tissue and others in unrelated tissues, suggesting that these pathologies may stem from shared molecular phenomena (**eTable 6**).

The mechanisms of action of many of the prognostic variants are currently unknown. There are many possibilities by which the variants that do not cause amino acid changes could affect cancer biology.^40^ Many variants are likely acting as *trans* eQTLs, which are difficult to study in datasets with relatively small sample sizes. The already high involvement of tumor suppressor genes, oncogenes, and driver genes among the prognostic germline variants is promising for future study. This report provides basic science researchers with genes and variants that should be studied to better understand the etiology and progression of cancers, while providing clinicians with the potential for better clinical predictions that could be made if germline variants are considered in the context of patient care.

## Data Availability

All of the raw data from The Cancer Genome Atlas used in this analysis is publicly available from the genomic data commons (https://portal.gdc.cancer.gov/).

## Acknowledgements

We thank Drs. Ana Damljanovic, Liz Williams, and Manisha Ray for their assistance with computation on the Cancer Genomics Cloud Platform, the High Performance Computing Team at the University of Virginia for assistance with computation on our university cluster and dbGAP for providing us with access to The Cancer Genome Atlas data. Most importantly, we are indebted to the patients and all of their families for their participation in The Cancer Genome Atlas project and the opportunity to study these cancers in a clinical context.

## Author Contributions

Conceptualization, A.C., A.D.; Methodology, A.C., A.R., A.D., X.T., T.L., P.K., M.K.; Software, Formal Analysis, Investigation, Writing – Original Draft, and Visualization, A.C.; Resources and Funding Acquisition, A.D., A.C.; Data Curation, A.C., R.P., Z.S., S.S., S.K., B.W., T.T., K.L., J.H.A., T.P., E.S., M.K.; Writing – Review & Editing, all authors; Supervision and Administration, A.D., A.R., P.K.

## Online Methods

### Data Sources, Variant Calling, and Quality Control

We determined the germline variant status of 10,582 cancer patients by variant calling the patients’ whole exome sequenced normal samples (WXS normal), whole exome sequenced tumor samples, (WXS tumor) and RNA sequenced tumor samples (RNA tumor) available on Cancer Genomics Cloud^41^ using VarDict (mapping quality > 30, base quality > 25, variant reads > 2, minimum allele frequency > 5%, no duplicate reads)^42^ and determined the sequencing depth at each position using samtools (mapping quality > 30).^43^ We set variant calls to unknown if the position at which the variant was called was covered by fewer than 10 reads. We then merged these three variant call sets, giving preference to WXS normal then WXS tumor then RNA tumor. We only included variants with an allele frequency of greater than 5% in the non-Finnish European population of gnomAD,^44^ variants found in more than 14 patients in a given cancer, and variants whose calls were greater than 90% concordant with each other in a given cancer in our final analysis. These thresholds had been selected in our previous study in order to better tune the allele frequencies of the European patients in our study to previously reported population frequencies.^30^ We labeled variant calls as concordant for a given variant if they gave the exact same variant call (homozygous for the reference allele, heterozygous, or homozygous for the alternate allele) in the WXS normal, WXS tumor, and RNA tumor samples. Variant calls were therefore discordant if the variant call differed in any of the three samples. The percentage concordance was calculated for each germline by dividing the total number of concordant variant calls by the total number of patients and multiplying the result by 100%.

We retrieved clinical outcomes data for each patient using the TCGA Pan-Cancer clinical data resource.^45^ We used TCGAbiolinks to obtain patient clinical information^46^ and we downloaded patient race composition from The Cancer Genome Ancestry Atlas (TCGAA).^47^ Additional clinical information for the lower grade glioma and glioblastoma patients was downloaded from a previous analysis.^48^ We used Lasso-regularization to determine which clinical covariates should be controlled for in our models, while using patient race composition from TCGAA in place of patient-reported race.^49,50^ We were not able to control for treatment. As discussed in greater detail by Liu et al., it is very difficult to control for treatment in the TCGA dataset.^45^ Detailed treatment information was not submitted in a consistent manner for many of the patients in TCGA and absence of submitted treatment information does not necessarily mean that the patient did not receive treatment. Furthermore, treatment regimens are quite complex and depend on chemotherapy drug selection and dosage, extent of surgical excision, and radiation therapy, among other factors. The broad spectrum of treatment options makes treatment challenging to control for. As discussed by Liu et al., the TCGA treatment information will likely need to be evaluated by panels of cancer specialists before it can be used for modeling in pan-cancer studies.^45^

We determined the number of somatic mutations in the cancer samples and evaluated the overlap between germline variants and somatic mutations and RNA editing sites as previously described.^30^ To ensure that our variant calls from the four variant call sets (WXS normal, WXS tumor, RNA tumor, and Combined) were concordant with each other, we calculated the allele frequency of each variant as in our previous analysis and calculated the Spearman correlation coefficient of these allele frequencies with each other.

### Power Analysis

We performed a power analysis in individual cancers to evaluate our ability to detect associations between germline variants and patient outcome using Cox regression. The power to detect an association between a germline variant and patient outcome is dependent on the sample size, effect size, correlation with other covariates in the model, the number of individuals with the germline variants, and the number of individuals without a germline variant, among other factors. As a result, the power to detect an association differs between germline variants, even assuming the same hazard ratio. To estimate our power, we therefore randomly sampled 10,000 germline for each cancer from the pool of germline variants to be tested in that cancer. We calculated statistical power using the powerSurvEpi R package (https://cran.r-project.org/web/packages/powerSurvEpi/index.html). We calculated our power to detect a significant association at a significance level (α) of:

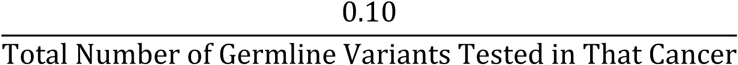

This threshold would be as stringent or slightly more stringent than false discovery correction using the Benjamini-Hochberg procedure which we ultimately used in our analysis. We then calculated the percentage of germline variants for which we had greater than 80% statistical power to detect a significant association at hazard ratios of 2, 3, 4, 5, 10, 15, and 20.

### Identification of Prognostic Germline Variants

We utilized six total approaches for identifying prognostic germline variants. In all analyses, we tested variants for an association with outcome using a Cox regression model, controlling for the covariates that we identified previously for each cancer using Lasso-regularization. We used the R packages survminer (https://cran.r-project.org/web/packages/survminer/index.html) and survival (https://cran.r-project.org/web/packages/survival/index.html) to perform Cox regression and generate Kaplan-Meier plots. p-values were corrected for multiple hypothesis testing using the Benjamini-Hochberg procedure. The circos plots were generated using the R package circlize.^51^

In analysis 1, we tested variants for an association with patient outcome in individual cancers, setting an adjusted p-value threshold (FDR) less than 0.10. We reported all statistically significant results and did not filter our results based on a hazard ratio threshold, as it is difficult to know what hazard ratio threshold would be clinically and biologically relevant. In the second analysis, we filtered our results from analysis one to identify germline variants that were recurrently associated (p<0.05) with favorable (hazard ratio (HR)<1) or poor (HR>1) outcome relative to the reference allele in seven or more cancers, such that the most recurrent prognostic variants would be reported. Given that molecular similarities between some of the TCGA cancers may have made it more likely that certain germline variants would be picked up in this second analysis than others, we did not think that it would be statistically valid to estimate the probability of variants being pulled out by this analysis by chance. In the third analysis, we grouped the cancers based on clinical understanding about the cancers and clustering patterns observed previously by the TCGA research network.^52^ We tested germline variants for associations with patient outcome (FDR<0.10) in these larger groups to detect germline variants with smaller effect sizes. In pooling cancers, we implicitly assumed that the germline variant had similar effects in the grouped cancers. If this assumption was not true for a particular germline variant, then that germline variant would actually be less likely to be associated with patient outcome. Only variants found in 15 or more patients across all grouped cancers were tested, resulting in fewer variants being tested in this analysis.

Analyses 4-6 were quite similar to analyses one through three, except that we restricted our analysis to only germline variants that caused significant amino acid changes with a combined annotation dependent depletion (CADD) score greater than 25.^53^ This enabled us to identify associations that we did not capture in analyses one through three due to the relatively higher stringency in that analysis resulting from multiple hypothesis correction. In analysis four, we tested variants with CADD score > 25 in individual cancers for an association with patient outcome (FDR<0.10). In analysis five, we filtered the results from analysis four to identify germline variants with CADD score > 25 that were recurrently associated (p<0.05) with favorable (HR<1) or poor (HR>1) prognosis in 5 or more patients. In analysis six, we tested germline variants with CADD > 25 for a significant association (FDR<0.10) with patient outcome in the previously described patient groups.

The Cox regression models that we fit for individual cancers controlled for the covariates that we found to be prognostic in those cancers (**eTable 1**). The Cox regression models that we fit for patient groups controlled for the covariates that we found to be prognostic in individual cancers with each term containing an interaction term associating that variable with the cancer that it was associated with patient outcome in. We also controlled for cancer type in these combined groups. As an example, suppose that variable A is associated with patient outcome in cancer X and variable B is associated with patient outcome in cancer Y. Then we would fit two Cox regression models to identify prognostic germline variants in individual cancers and a third Cox regression model to identify germline variants prognostic in the pooled cohort, as illustrated below.

1. Identifying germline variants associated with patient outcome in cancer X

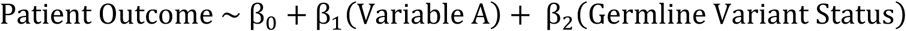
2. Identifying germline variants associated with patient outcome in cancer Y

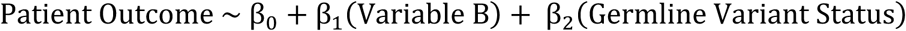
3. Identifying germline variants associated with patient outcome when the patients with cancer X and the patients with cancer Y are pooled together

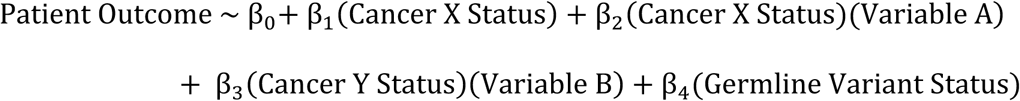

In model (3) above, cancer X status is a dummy variable that can be 0 or 1. The value of this variable is 0 for patients with cancer Y and 1 for patients with cancer X. The opposite is true for the cancer Y status variable. This allowed us to group patients to test for an association with patient outcome, while controlling for differences between different cancers and relevant clinical differences between patients with the same cancer.

### Concordance and Correlation of Hazard Ratios for the Prognostic Germline Variants

We tested whether germline variants associated with patient outcome (p<0.05) in three of more cancers were typically recurrently deleterious or protective more often than would be expected by random chance and if the hazard ratios estimated for these prognostic germline variants in different cancers were correlated with each other.

To test for concordance, we first counted the number of times that germline variant was found to be associated (p<0.05) with poor patient outcome (HR<1) or favorable patient outcome (HR>1). We then calculated the following value for each prognostic germline variant:

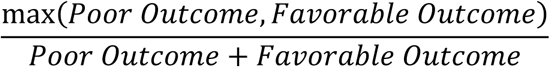

where poor outcome is the number of times that the germline variant was associated with poor outcome (HR<1) and favorable outcome is the number of times that the germline variant was associated with favorable outcome (HR>1). If a germline variant was perfectly concordant, then the calculated value would be 1. While theoretically the expected value would be 0.5 for a random germline variant, we empirically estimated the expected value by the following calculation:

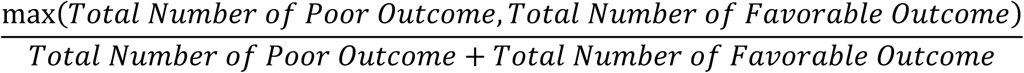

In this set of prognostic variants, there were more variants associated with poor patient outcome (HR<1) than favorable patient outcome (HR>1), resulting in the expected index being 0.589. We then used a Wilcoxon rank sum test to determine whether the concordance values that we calculated from the set of prognostic germline variants differed from what we would expect by random chance.

We next tested whether the hazard ratios estimated for a given prognostic germline variant in different cancers were correlated with each other. Because we had previously found the hazard ratios to be concordant, we performed this analysis separately for instances in which a germline variant was found to be deleterious and protective. We identified the set of variants associated with favorable (HR<1) outcome and deleterious (HR>1) outcome in three or more cancers. The set of variants that were associated with favorable and deleterious outcome were analyzed separately. For each analysis, we generated all possible pairs of hazard ratios for a given germline variant. We then ran a Spearman’s correlation test to determine whether or not the hazard ratios were correlated to each other. Because the hazard ratio is also correlated to the allele frequency, we repeated the prior analysis with a Spearman partial correlation test to control for germline variant allele frequency. Partial correlation was calculated used the ppcor R package.^54^

### Characteristics of Prognostic Germline Variants

Having identified the prognostic germline variants, we then aimed to compare the characteristics of prognostic germline variants to the characteristics of germline variants identified in previous genome wide association studies.^31^ We decided to use the variants from analysis one and analysis three to understand the characteristics of prognostic germline variants because the other approaches each identified a very small number of prognostic germline variants. We decided not to pool all of the germline variants together due to possible differences in characteristics between these sets of variants. We therefore analyzed the characteristics of the prognostic germline variants from analysis one and from analysis three separately. To avoid considering the same information multiple times, we removed variants that were linked with each other from the analyses in this section and only retained the first variant by genomic position. The actual variant retained did not have a significant effect on our results because the hazard ratios and sample sizes for the linked variants were very similar.

We first tested whether or not the minor allele was typically associated with poor patient outcomes. We sorted the variants into two categories: minor alleles that were deleterious in the Cox regression model (HR>1) and minor alleles that were associated with favorable outcomes (HR<1). Although the reference allele was often the major allele, this was not always the case. We performed a one-sided Fisher’s exact test in R to determine whether or not the minor allele was more likely to be deleterious. The R package scatterpie (https://cran.r-project.org/web/packages/scatterpie/index.html) was used to display the proportion of homozygous reference, heterozygous, and homozygous alternate individuals in **Figures 4A-B**. For variants in analysis three that were pulled out in multiple groups, we displayed the proportion of individuals only for the group that contained the largest number of individuals. The largest group always contained all individuals because the smaller groups were made up of smaller number of cancers and was always contained in the larger group. For example, suppose a variant was found to be prognostic in both group 20 (KICH, KIRP) and group 19 (KICH, KIRC, KIRP). In this case, we would perform all calculations using the information from group 19.

We next tested whether or not there was an inverse correlation between effect size and allele frequency. To do this, we calculated the Spearman correlation coefficient between effect size, calculated as |ln(HR) − 0|, and allele frequency. Finally, we identified the genomic regions (upstream of a gene, 5’ UTR, exonic, intronic, 3’ UTR, downstream of a gene, or intergenic) in which each variant was located in using annovar.^55^ Some variants were found in multiple different transcripts and therefore mapped to several different genic regions. For the purposes of creating **Figures 4C-D**, we allowed a single variant to count once for multiple different regions. Excluding these variants from **Figures 4C-D** did not change our interpretation of the results.

### Association of Prognostic Germline Variants with Somatic Driver Mutations

We tested whether the prognostic germline variants were more likely to be associated with somatic mutations in driver genes than would be expected by random chance. We retrieved the set of driver genes for each cancer and consensus somatic mutation calls for each cancer from TCGA Network analyses.^2,56^ For each cancer, we only considered driver genes with five or more patients with a somatic mutation in that driver gene in that cancer. For each prognostic germline variant, we tested whether the deleterious variant was associated with an increased incidence of somatic mutations in each of the driver genes being considered for that cancer in patients with the deleterious allele compared to patients with the protective allele using a one-sided Fisher’s exact test. p-values were adjusted using the Benjamini-Hochberg procedure.

We were then able to determine the number of germline variants that were associated with a somatic mutation in a driver gene. We repeated this approach for all germline variants included in this analysis and performed a one-sided Fisher’s exact test to determine whether or not more prognostic germline variants than expected were associated with a somatic mutation in a driver gene.

### Area Under the Curve

To assess the clinical relevance of our findings, we tested whether the germline variants enhanced patient outcome predictions made using clinical information alone. While we had identified germline variants associated with outcome controlling for clinical covariates, we aimed to determine whether these variants significantly improved patient outcome predictions beyond predictions made using the clinical model alone, particularly in cancers in which the prediction by the clinical model was already quite accurate. We generated receiver operator characteristic (ROC) curves from the tenth percentile of patient death or patient progression to the ninetieth percentile of patient death or patient progression for each variant in R (https://cran.r-project.org/web/packages/survivalROC/survivalROC.pdf, https://cran.r-project.org/web/packages/timeROC/timeROC.pdf). We generated two ROC curves per variant: (1) the first was made using only patient clinical information (C) and (2) the second was generated using both patient clinical information and germline variant status (C+GV). We ran a one-sided Wilcoxon-rank sum test in R to determine whether the model supplemented with germline variant status consistently yielded better predictions across time for each variant. While our Cox regression analysis identified variants that were significantly associated with patient outcome, these variants may not necessarily substantially improve clinical outcome predictions in cancers in which the clinical variables are already very good at predicting outcome. Running the one-sided Wilcoxon-rank sum test allowed us to test whether the improvement to the prediction was significant.

### Gene Annotation and Literature Review

We annotated the variants resulting from our analysis using biomaRt.^57,58^ We reviewed the literature for the functions of these genes to understand their functions. Many of the authors (RP, SK, ZS, SS, BW, TT, JA, KL, TP, ES, MK) initially reviewed the literature for information about each gene. The literature review was then verified by three of the authors (RP, SK, ZS) to ensure consistency and validity.

Having generated a list of genes that the germline variants are associated with from biomaRt, we first specifically searched the literature to see if these genes had a function in cancer that had been characterized and that fit a category described by Weinberg and Hanahan.^32^ This part of the literature review had the largest number of unknowns due to the large amount of specificity required by the studies. We then relaxed our stringency and checked to see whether or not the gene was associated with findings in the literature consistent with oncogenic or tumor suppressor activity in the context of cancer. Finally, to understand in general whether or not these genes are being actively studied by the field, we categorized these genes based on whether or not the literature suggested that the genes are being studied in a cancer in which the germline variant was found to be prognostic, studied in any cancer, or studied in any human disease. We also overlapped our gene list with the list of driver genes generated by the TCGA research network.^2^

### Variant Mechanisms and Literature Review

We next aimed to understand the mechanisms by which the prognostic germline variants may be exerting their effects. We started with the germline variants that were predicted to cause significant amino acid changes (CADD>25). We determined the position and amino acid change caused by these germline variants using Ensembl.^59^ We determined the domain in which these germline variants cause their amino acid changes using the National Center for Biotechnology Information databases (https://www.ncbi.nlm.nih.gov/) and the Ensembl and Uniprot^60^ databases. We next identified germline variants that are likely acting as expression quantitative trait loci in *cis* (*cis* eQTLs). For each germline variant, we separated patients based on whether or not they had at least one non-reference allele and then determined whether or not there was a statistically significant difference between the mean expression of the gene associated with the variant between the two groups using a Wilcoxon rank sum test. We then combined our prediction as to whether the germline variant was protective or deleterious with the expression difference between the two groups to determine whether increased expression of the gene would be expected to be protective or deleterious. We fit Cox regression models using the expression of each of the genes, controlling for clinical covariates, and compared the result to our prediction. We reported variants that are concordant with our predictions. Because the differential expression and Cox regression results had to both be concordant with each other, we used a more relaxed cut-off of p < 0.10 for hypothesis generation. Further studies with larger cohorts and more statistically power are necessary to further interrogate these associations. Finally, we checked to see whether the eQTL was also reported in GTEx in the tissue from which the tumor was derived by downloading the list of tissue-specific and pan-tissue eQTLs and comparing the eQTLs identified in our analysis to those reported in GTEx.

We reviewed the literature for previous associations tied to these variants reported in the literature. As was the case with gene annotation, the literature review was first done by multiple authors (RP, SK, ZS, SS, BW, TT, JA, KL, TP, ES, MK) with the final round of quality control and verification being done by a single author (BW).

### Correlation with Drug Sensitivity

We found the germline variant rs1800932 in *MSH6* to be associated with favorable patient outcome and increased *MSH6* expression. Because a previous analysis found that *MSH6* knockdown resulted in increased temozolomide resistance,^36^ we tested whether *MSH6* expression was correlated with temozolomide sensitivity in cancer cell lines. To do this, we downloaded *MSH6* expression levels and temozolomide sensitivity for 915 cell lines using data from the Genomics of Drug Sensitivity in Cancer database^61^ through CellMinerCDB.^35^ We tested for an association using Spearman’s correlation test.

### Pathway Dysregulation

For selected prognostic germline variants described in the text, we tested whether or not these prognostic germline variants were associated with upregulation or downregulation of genes in specific pathways. For each prognostic germline variant, we separated patients into two groups based on whether or not the variant allele was called in those patients. We calculated the log fold change of each gene expressed greater than a median of 1 fragment per kilobase per million mapped reads and used these values as an input for gene set enrichment analysis.^62^

## References

1. Hudson, T.J., et al. International network of cancer genome projects. Nature 464, 993–998 (2010).

2. Bailey, M.H., et al. Comprehensive Characterization of Cancer Driver Genes and Mutations. Cell 173, 371-385.e318 (2018).

3. Lee, B., et al. Exploring the feasibility and utility of exome-scale tumour sequencing in a clinical setting. Internal medicine journal 48, 786–794 (2018).

4. Lichtenstein, P., et al. Environmental and heritable factors in the causation of cancer--analyses of cohorts of twins from Sweden, Denmark, and Finland. The New England journal of medicine 343, 78–85 (2000).

5. Huang, K.L., et al. Pathogenic Germline Variants in 10,389 Adult Cancers. Cell 173, 355-370.e314 (2018).

6. Zhang, J., et al. Germline Mutations in Predisposition Genes in Pediatric Cancer. The New England journal of medicine 373, 2336–2346 (2015).

7. Pearlman, R., et al. Prevalence and Spectrum of Germline Cancer Susceptibility Gene Mutations Among Patients With Early-Onset Colorectal Cancer. JAMA Oncol 3, 464–471 (2017).

8. Mandelker, D., et al. Mutation Detection in Patients With Advanced Cancer by Universal Sequencing of Cancer-Related Genes in Tumor and Normal DNA vs Guideline-Based Germline Testing. Jama 318, 825–835 (2017).

9. Cheng, D.T., et al. Comprehensive detection of germline variants by MSK- IMPACT, a clinical diagnostic platform for solid tumor molecular oncology and concurrent cancer predisposition testing. BMC Med Genomics 10, 33 (2017).

10. Lee, S.E., et al. High prevalence of the MLH1 V384D germline mutation in patients with HER2-positive luminal B breast cancer. Sci Rep 9, 10966 (2019).

11. Shivakumar, M., Miller, J.E., Dasari, V.R., Gogoi, R. & Kim, D. Exome-Wide Rare Variant Analysis From the DiscovEHR Study Identifies Novel Candidate Predisposition Genes for Endometrial Cancer. Front Oncol 9, 574 (2019).

12. Gori, S., et al. Recommendations for the implementation of BRCA testing in ovarian cancer patients and their relatives. Crit Rev Oncol Hematol 140, 67–72 (2019).

13. Tian, W., et al. Screening for hereditary cancers in patients with endometrial cancer reveals a high frequency of germline mutations in cancer predisposition genes. Int J Cancer 145, 1290–1298 (2019).

14. Menden, M.P., et al. The germline genetic component of drug sensitivity in cancer cell lines. Nature communications 9, 3385 (2018).

15. Pomerantz, M.M., et al. The association between germline BRCA2 variants and sensitivity to platinum-based chemotherapy among men with metastatic prostate cancer. Cancer 123, 3532–3539 (2017).

16. Low, S.K., Zembutsu, H. & Nakamura, Y. Breast cancer: The translation of big genomic data to cancer precision medicine. Cancer Sci 109, 497–506 (2018).

17. Hahnen, E., et al. Germline Mutation Status, Pathological Complete Response, and Disease-Free Survival in Triple-Negative Breast Cancer: Secondary Analysis of the GeparSixto Randomized Clinical Trial. JAMA Oncol 3, 1378–1385 (2017).

18. Li, X., Wu, N. & Li, B. A high mutation rate of immunoglobulin heavy chain variable region gene associates with a poor survival and chemotherapy response of mantle cell lymphoma patients. Medicine 98, e15811 (2019).

19. Horak, P., et al. Response to olaparib in a PALB2 germline mutated prostate cancer and genetic events associated with resistance. Cold Spring Harb Mol Case Stud 5(2019).

20. Crona, D.J., et al. Genetic Variants of VEGFA and FLT4 Are Determinants of Survival in Renal Cell Carcinoma Patients Treated with Sorafenib. Cancer research 79, 231–241 (2019).

21. de Velasco, G., et al. Pharmacogenomic Markers of Targeted Therapy Toxicity in Patients with Metastatic Renal Cell Carcinoma. European urology focus 2, 633–639 (2016).

22. Hertz, D.L., Henry, N.L. & Rae, J.M. Germline genetic predictors of aromatase inhibitor concentrations, estrogen suppression and drug efficacy and toxicity in breast cancer patients. Pharmacogenomics 18, 481–499 (2017).

23. Lee, S.H.R. & Yang, J.J. Pharmacogenomics in acute lymphoblastic leukemia. Best Pract Res Clin Haematol 30, 229–236 (2017).

24. Singh, M., Bhatia, P., Khera, S. & Trehan, A. Emerging role of NUDT15 polymorphisms in 6-mercaptopurine metabolism and dose related toxicity in acute lymphoblastic leukaemia. Leuk Res 62, 17–22 (2017).

25. Guan, J., et al. Clinical response of the novel activating ALK-I1171T mutation in neuroblastoma to the ALK inhibitor ceritinib. Cold Spring Harb Mol Case Stud 4(2018).

26. Udagawa, C., et al. Whole exome sequencing to identify genetic markers for trastuzumab-induced cardiotoxicity. Cancer Sci 109, 446–452 (2018).

27. Carter, H., et al. Interaction Landscape of Inherited Polymorphisms with Somatic Events in Cancer. Cancer discovery 7, 410–423 (2017).

28. Guerrini-Rousseau, L., et al. Germline SUFU mutation carriers and medulloblastoma: clinical characteristics, cancer risk, and prognosis. Neurooncology 20, 1122–1132 (2018).

29. Baretta, Z., Mocellin, S., Goldin, E., Olopade, O.I. & Huo, D. Effect of BRCA germline mutations on breast cancer prognosis: A systematic review and meta-analysis. Medicine 95, e4975 (2016).

30. Chatrath, A., Kiran, M., Kumar, P., Ratan, A. & Dutta, A. The Germline Variants rs61757955 and rs34988193 are Predictive of Survival in Lower Grade Glioma Patients. Molecular cancer research : MCR (2019).

31. Park, J.H., et al. Distribution of allele frequencies and effect sizes and their interrelationships for common genetic susceptibility variants. Proceedings of the National Academy of Sciences of the United States of America 108, 18026–18031 (2011).

32. Hanahan, D. & Weinberg, R.A. Hallmarks of cancer: the next generation. Cell 144, 646–674 (2011).

33. GTEx-Consortium. The Genotype-Tissue Expression (GTEx) project. Nature genetics 45, 580–585 (2013).

34. Basu, S., et al. A study of molecular signals deregulating mismatch repair genes in prostate cancer compared to benign prostatic hyperplasia. PloS one 10, e0125560 (2015).

35. Rajapakse, V.N., et al. CellMinerCDB for Integrative Cross-Database Genomics and Pharmacogenomics Analyses of Cancer Cell Lines. iScience 10, 247–264 (2018).

36. McFaline-Figueroa, J.L., et al. Minor Changes in Expression of the Mismatch Repair Protein MSH2 Exert a Major Impact on Glioblastoma Response to Temozolomide. Cancer research 75, 3127–3138 (2015).

37. Xie, C., et al. Association of MSH6 mutation with glioma susceptibility, drug resistance and progression. Molecular and clinical oncology 5, 236–240 (2016).

38. Baldari, S., Ubertini, V., Garufi, A., D’Orazi, G. & Bossi, G. Targeting MKK3 as a novel anticancer strategy: molecular mechanisms and therapeutical implications. Cell death & disease 6, e1621 (2015).

39. Ding, L., et al. Perspective on Oncogenic Processes at the End of the Beginning of Cancer Genomics. Cell 173, 305-320.e310 (2018).

40. Zhang, F. & Lupski, J.R. Non-coding genetic variants in human disease. Hum Mol Genet 24, R102–110 (2015).

41. Lau, J.W., et al. The Cancer Genomics Cloud: Collaborative, Reproducible, and Democratized-A New Paradigm in Large-Scale Computational Research. Cancer research 77, e3–e6 (2017).

42. Lai, Z., et al. VarDict: a novel and versatile variant caller for next-generation sequencing in cancer research. Nucleic acids research 44, e108 (2016).

43. Li, H., et al. The Sequence Alignment/Map format and SAMtools. Bioinformatics (Oxford, England) 25, 2078–2079 (2009).

44. Lek, M., et al. Analysis of protein-coding genetic variation in 60,706 humans. Nature 536, 285–291 (2016).

45. Liu, J., et al. An Integrated TCGA Pan-Cancer Clinical Data Resource to Drive High-Quality Survival Outcome Analytics. Cell 173, 400-416.e411 (2018).

46. Colaprico, A., et al. TCGAbiolinks: an R/Bioconductor package for integrative analysis of TCGA data. Nucleic acids research 44, e71 (2016).

47. Yuan, J., et al. Integrated Analysis of Genetic Ancestry and Genomic Alterations across Cancers. Cancer cell 34, 549-560.e549 (2018).

48. Ceccarelli, M., et al. Molecular Profiling Reveals Biologically Discrete Subsets and Pathways of Progression in Diffuse Glioma. Cell 164, 550–563 (2016).

49. Tibshirani, R. The lasso method for variable selection in the Cox model. Stat Med 16, 385–395 (1997).

50. Friedman, J., Hastie, T. & Tibshirani, R. Regularization Paths for Generalized Linear Models via Coordinate Descent. Journal of statistical software 33, 1–22 (2010).

51. Gu, Z., Gu, L., Eils, R., Schlesner, M. & Brors, B. circlize Implements and enhances circular visualization in R. Bioinformatics (Oxford, England) 30, 2811–2812 (2014).

52. Hoadley, K.A., et al. Cell-of-Origin Patterns Dominate the Molecular Classification of 10,000 Tumors from 33 Types of Cancer. Cell 173, 291- 304.e296 (2018).

53. Rentzsch, P., Witten, D., Cooper, G.M., Shendure, J. & Kircher, M. CADD: predicting the deleteriousness of variants throughout the human genome. Nucleic acids research 47, D886–d894 (2019).

54. Kim, S. ppcor: An R Package for a Fast Calculation to Semi-partial Correlation Coefficients. Communications for statistical applications and methods 22, 665–674 (2015).

55. Wang, K., Li, M. & Hakonarson, H. ANNOVAR: functional annotation of genetic variants from high-throughput sequencing data. Nucleic acids research 38, e164 (2010).

56. Ellrott, K., et al. Scalable Open Science Approach for Mutation Calling of Tumor Exomes Using Multiple Genomic Pipelines. Cell systems 6, 271- 281.e277 (2018).

57. Durinck, S., et al. BioMart and Bioconductor: a powerful link between biological databases and microarray data analysis. Bioinformatics (Oxford, England) 21, 3439–3440 (2005).

58. Durinck, S., Spellman, P.T., Birney, E. & Huber, W. Mapping identifiers for the integration of genomic datasets with the R/Bioconductor package biomaRt. Nature protocols 4, 1184–1191 (2009).

59. Cunningham, F., et al. Ensembl 2019. Nucleic acids research 47, D745–d751 (2019).

60. Consortium, U. UniProt: a worldwide hub of protein knowledge. Nucleic acids research 47, D506–d515 (2019).

61. Yang, W., et al. Genomics of Drug Sensitivity in Cancer (GDSC): a resource for therapeutic biomarker discovery in cancer cells. Nucleic acids research 41, D955–961 (2013).

62. Subramanian, A., et al. Gene set enrichment analysis: a knowledge-based approach for interpreting genome-wide expression profiles. Proceedings of the National Academy of Sciences of the United States of America 102, 15545–15550 (2005).

